# An intragenic duplication within *SIRPβ1* shows a dual effect over Alzheimer’s disease cognitive decline altering the microglial response

**DOI:** 10.1101/2022.11.19.22282342

**Authors:** José María García-Alberca, Itziar de Rojas, Elisabeth Sanchez-Mejias, Diego Garrido-Martín, Laura Gonzalez-Palma, Sebastian Jimenez, Almudena Pino-Angeles, Jose Manuel Cruz-Gamero, Silvia Mendoza, Emilio Alarcón-Martín, The GERALD consortium, Clara Muñoz-Castro, Luis Miguel Real, Juan Jesus Tena, Rocio Polvillo, Fernando Govantes, Aroa Lopez, Jose Luis Royo-Aguado, Victoria Navarro, Irene Gonzalez, Maximiliano Ruiz, Armando Reyes-Engel, Esther Gris, Maria Jose Bravo, Lidia Lopez-Gutierrez, Marina Mejias-Ortega, Paz De la Guía, María López de la Rica, Olga Ocejo, Javier Torrecilla, Carmen Zafra, María Dolores Nieto, Concepción Urbano, Rocío Jiménez-Sánchez, Nuria Pareja, Macarena Luque, María García-Peralta, Rosario Carrillejo, María del Carmen Furniet, Lourdes Rueda, Ana Sánchez-Fernández, Tomás Mancilla, Isabel Peña, Natalia García-Casares, Sonia Moreno-Grau, Isabel Hernández, Laura Montrreal, Inés Quintela, Antonio González-Pérez, Miguel Calero, Emilio Franco-Macías, Juan Macías, Manuel Menéndez-González, Ana Frank-García, Raquel Huerto Vilas, Mónica Diez-Fairen, Carmen Lage, Sebastián García-Madrona, Pablo García-González, Sergi Valero, Oscar Sotolongo-Grau, Alba Pérez-Cordón, Alberto Rábano, Alfonso Arias Pastor, Ana Belén Pastor, Ana Espinosa, Anaïs Corma-Gómez, Ángel Martín Montes, Ángela Sanabria, Carmen Martínez Rodríguez, Dolores Buiza-Rueda, Eloy Rodriguez-Rodriguez, Gemma Ortega, Ignacio Alvarez, Irene Rosas Allende, Juan A Pineda, Maitée Rosende-Roca, María Bernal Sánchez-Arjona, Marta Fernández-Fuertes, Montserrat Alegret, Natalia Roberto, Teodoro del Ser, Guillermo Garcia-Ribas, Pascual Sánchez-Juan, Pau Pastor, Gerard Piñol-Ripoll, María José Bullido, Victoria Álvarez, Pablo Mir, Miguel Medina, Marta Marquié, María Eugenia Sáez, Ángel Carracedo, Marina Laplana, Laura Tomas-Gallardo, Adelina Orellana, Lluís Tárraga, Mercè Boada, Joan Fibla Palazon, Javier Vitorica, Agustín Ruiz, Roderic Guigo, Antonia Gutierrez, Jose Luis Royo

## Abstract

Microglia play an important role in the maintenance of brain homeostasis, and microglial dysfunction plays a causative role in Alzheimer disease pathogenesis. Here we focus on the signal regulatory protein SIRPβ1, a surface receptor expressed on the myeloid cells that triggers amyloid-β and cell debris phagocytosis via TYROBP. We found that a common intragenic duplication alters the SIRPβ1 protein isoform landscape affecting both extracellular and transmembrane domains, which compromise their ability to bind oligomeric Aβ and their affinity for TYROBP. Epidemiological studies show that patients with mild cognitive impairment that are homozygous for the *SIRPβ1* duplication allele show an increased cerebrospinal fluid t-Tau/Aβ ratio (p-value=0.018) and a higher risk to develop AD (OR=1.678, p-value=0.018). Magnetic resonance imaging at diagnosis showed that AD patients with the duplication allele exhibited a worse initial response to the disease. At the moment of diagnosis all patients showed equivalent Mini-Mental State Examination scores. However AD patients with the duplication allele had less hippocampal degeneration (Beta= -0.62, p-value < 0.001) and fewer white matter hyperintensities. In contrast, longitudinal studies indicate that patients bearing the duplication allele show a slower cognitive decline after correcting by baseline (p-value = 0.013). Transcriptional analysis of the patients’ hippocampus also shows that the *SIRPβ1* duplication allele correlates with higher *TREM2* expression and an increased microglial activation. Given the recent pharmacological approaches focused on the TREM2-TYROBP axis, we consider that the presence of this structural variant might be considered as a potential modulator of this causative pathway.

## Introduction

Alzheimer_’_s Disease (AD) is characterized by the formation of extracellular plaques containing amyloid-β (Aβ) aggregates^1^. Microglia, the brain-resident macrophages, represent the largest population of myeloid cells in the brain parenchyma^2^. They quickly respond and surround amyloid plaques acquiring a disease-associated state. This is characterized by downregulation of homeostatic genes and upregulation of disease-associated genes including apolipoprotein E (*APOE*) and triggering receptor expressed on myeloid cells 2 (*TREM2*), the two major genetic risk factors for late-onset AD. The identification of microglial AD risk gene variants supports a causative role for these cells in the disease^3,4^. Microglial activation is tightly regulated by molecular mechanisms that are promptly switched on as the first responders to noxious stimuli, and rapidly turned off to avoid unwanted effects. Signal Induced Proteins (SIRPs) are a membrane protein family expressed in myeloid cells including microglia. They share the typical structural organization of the immunoglobulin superfamily with an N-terminal extracellular domain containing three cysteine-bound Ig-like loops and a single membrane-spanning transmembrane domain (TMD)^5^. The C-terminal intracellular domains of the SIRPα subfamily contain a relatively long amino acid sequence that includes four tyrosine residues forming two immuno-receptor tyrosine-based inhibition motifs. Conversely, the SIRPβ subfamily members have a short intracellular domain containing only a few amino acids with no reported activity. Previous data demonstrated that SIRPβ1 TMD mediates interactions with the transmembrane immune signaling adaptor TYROBP, which has an intracellular immuno-receptor tyrosine-based activation motif^6^. Peripherally, SIRPβ1 positively regulates macrophage phagocytosis^7^ and neutrophil migration^8^. In the central nervous system (CNS), SIRPβ1 is upregulated in microglia of APP transgenic mice as well as in AD patients. Activation of SIRPβ1 induces reorganization of cytoskeletal proteins, counterregulates proinflammatory mediators, and increases microglial phagocytosis of microsphere beads, neural debris, and fibrillary Aβ. In contrast, lentiviral knockdown of SIRPβ1 impaired microglial phagocytosis of neural cell debris and Aβ^9^.These findings support the hypothesis that SIRPβ1-mediated signal triggers microglial response.

TYROBP signaling has been proposed to be the main regulator in the transformation process from a homeostatic to a disease-associated state in microglia^10^. Mutations in the TYROBP main ligand, TREM2, are strong risk factors for AD^11 12,13^. The TREM2-TYROBP axis has also been shown to control microglial activity and consequently affect the fate of damaged neurons after neuronal injury ^14,15^. In fact, among the signaling pathways required for maintaining correct neuron-microglia relationships, the TREM2-TYROBP axis is key. TREM2 signaling is essential for maintaining CNS tissue homeostasis. TREM2-TYROBP-mediated phagocytosis of accidental apoptotic materials is a beneficial function of microglia that takes place without frank inflammation. However, excessive neuronal cell death could generate extensive inflammatory processes with deleterious effects on the brain. Different TREM2-positive phenotypes can be assumed by microglia depending on local and temporal conditions, with or without inflammation, generating protective or detrimental effects ^16^.

We have characterized a common Copy-Number Variant (CNV) consisting of a 30Kb insertion within the *SIRPβ1 locus* derived from an ancestral duplication^17^,^18^. Recent data have revealed its role in systemic immunity because of its link to cancer aggressiveness and viral infection susceptibility ^19,20^. Here, we explore whether this *SIRPβ1* structural variant might jeopardize the TREM2-TYROBP axis and affect the microglial activity of AD patients. We have found that the *SIRPβ1* duplication allele leads to the appearance of isoforms with alternative extracellular structures and TMDs with differential affinity for TYROBP. This results in uneven abilities to bind Aβ and induce the phagocytosis *in vitro*. Our results also show that mild cognitive impairment patients with the *SIRPβ1* duplication have a higher risk to develop AD and display a worse initial response to the disease. However, as the disease advances, the *SIRPβ1* duplication allele is associated with a slower cognitive decline, suggesting a dual effect along the disease course.

## Results

### *SIRPβ1* duplication increases the risk of dementia among mild cognitive patients while delaying cognitive progression of AD patients

The interstitial *SIRPβ1* CNV consists of an ancestral duplication nowadays observed as a common insertion with an allele frequency of ≈0.2 in the general population. Previous reports estimated an r^2^=0.98 between the single nucleotide polymorphism rs2209313 allele T and the duplication allele of the *SIRPβ1* CNV^17, 21^. We independently confirmed these calculations taking advantage of the 1,000 Genomes Project data (Extended Methods) and therefore rs2209313 has been genotyped as a proxy of this structural variant. In order to explore whether this *locus* might affect AD risk, we performed a case-control association meta-analysis using the available datasets together with our own data from the DEGESCO consortium^22^. The combined results show that the rs2209313-T (duplication) allele has no significant effect over AD risk *per se* (OR_meta_=0.980, p=0.06, Extended Data Table 1). Next, we explored the conversion risk from mild cognitive impairment (MCI) to AD, determined in the GR@ACE series, a subcohort of DEGESCO consortium which comprised 1,615 MCI patients who were followed up. We observed that MCI patients homozygous for the rs2209313 allele T-duplication (hereafter Dup/Dup) showed an increased likelihood to develop AD (Extended Data Table 2). Cerebrospinal fluid (CSF) taken at the moment of MCI diagnosis was analyzed, and we could observe that Dup/Dup patients showed increased phospho and total Tau/Aβ_42_ ratios (Extended Figure 1). This coincides with the epidemiological data of a worse prognosis of MCI patients. To gain further insight into this effect, we took advantage of the GERALD series, another subcohort of DEGESCO which has magnetic resonance imaging (MRI) data taken at the moment of AD diagnosis. Patients homozygous for the duplication display less medial temporal atrophy (Scheltens visual ranking average of 4.5 *vs* 0.8) than those homozygous for the rs2209313 allele C-Wt (hereafter Wt/Wt) after adjusting by ApoE and hypertension (β = -0.605, p-value < 0.001) (Fig. 1a, 1b). Equivalent results were obtained with ANCOVA analysis (F_2,99_ = 32.402, p-value < 0.001). In the same way, Dup/Dup patients show fewer White Matter Hyperintensities (WMH) than Wt/Wt patients after the same adjustments (Scheltens visual ranking average of 3.38 *vs* 1.0 for Total Periventricular Hyperintensities –TPVH–) (F_2, 99_ = 10.148, p-value = 0.044), suggesting lower Aβ_42_-associated vascular damage (Fig. 1a, 1c). Since WMH is associated with hypertension, we split the GERALD series according to the hypertensive status (Extended Data Fig. 2). The results suggest that for both independent subgroups, Dup/Dup patients showed lower levels of vascular damage. The Mini-Mental State Examination (MMSE) scores were similar among the three groups (Figure 1d), suggesting that at the moment of AD diagnosis, the Dup/Dup patients presented equivalent cognitive functions with significantly less neural damage. This associates the *SIRPβ1* Dup allele with a worse first-response to the initial stress of the CNS. We next examined whether this effect had a clinical correlation during the disease course. To this end, we conducted a prospective study evaluating the cognitive progression in the patients included in the GERALD series, and calculated the slope between the MMSE scores obtained upon recruitment and the first follow-up (FFU) (Extended Data Table 1). GERALD patients were then stratified according to their cognitive decline. Patients with a cognitive slope below the median were considered as slow progressors in contrast to the rapid ones. Binary logistic regression showed that AD patients bearing the *SIRPβ1* Dup/Dup genotype also displayed a slower cognitive decline than the wild-type genotype after adjusting by *APOE* and MMSE score at diagnosis (Wald = 4.084, OR = 2.989, p-value = 0.043). Considering the entire series and stratifying them according to the *SIRPβ1* genotype, we observed that Dup/Dup patients showed a slower cognitive decline expressed in MMSE scores month^-1^ (average ± SE: -0.09 ± 0.09, n = 10) than the Wt/Wt patients (−0.13 ± 03, n = 106). Linear regression did not reach statistical significance when adjusting by MMSE baseline and *APOE* genotype (Fig. 1e). Next, we replicated our longitudinal analysis in the GR@ACE cohort, an independent series which accounts with 4,656 AD patients, being the genetic effect statistically significant (β= 0.037, p-value = 0.013) (Fig. 1f). We should highlight this dual effect on the Dup/Dup genotype. First, the mutant allele was found to be a risk factor among MCI patients with a higher risk to convert AD. It was also associated with worse CSF biomarker levels and at a structural level, lower MTA atrophy and vascular damage generated the same loss of MMSE score than the Wt allele. In contrast, the duplication allele showed a protective effect over the cognitive decline in two independent series, suggesting a higher level of resilience of the CNS once the disease progresses. However, the best fitting genetic model in the case of the GR@ACE series was recessive rather than the codominant one observed in the GERALD series. One particular aspect that may explain this apparent discrepancy between both series is the difference in intervals of follow-up assessments. In the GR@ACE series, patients had their second visit 10.2 months after diagnosis, whereas in the GERALD it was at 18.2 months (Extended Data Table 3). Therefore, our independent observations might indeed be the same effect at two different timepoints (Fig. 1g).

**Figure 1.**
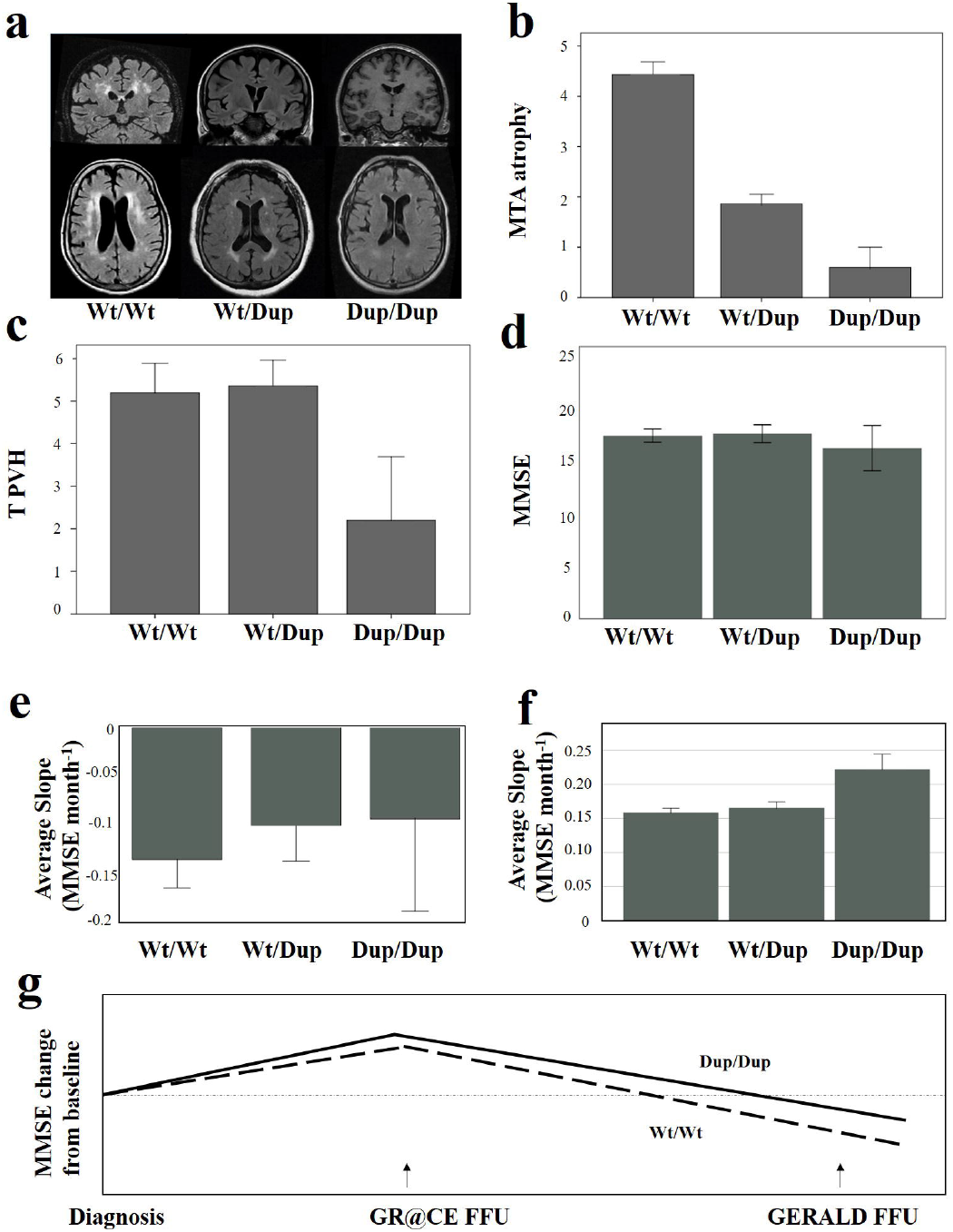
Impact of *SIRPβ1* rs2209313 in brain damage in AD patients. Impact of rs2209313 allele T-duplication in AD evolution. **a**, coronal (upper row) view of MTA from three illustrative patients with the three rs2209313 genotypes and sagittal (lower row) WHM from three illustrative patients with the different possible genotypes. **b**, total periventricular hyperintensities from the Wt/Wt, n = 89; Wt/Dup, n = 51 and Dup/Dup, n=8 patients. **c**, medial temporal atrophy from the same patients. **d**, MMSE scores obtained at the moment of diagnosis. **e**, average MMSE slope values calculated as points of MMSE changed per month between the diagnosis and the first follow up (FFU) for the patients of GERALD (Wt/Wt, n = 106; Wt/Dup, n = 65 and Dup/Dup, n = 10) and **f**, GR@ACE series (Wt/Wt, n = 3,037; Wt/Dup, n = 1,429 and Dup/Dup, n = 190). **g**, effect of the mutation over the cognitive decline observed in both series, given that the GR@ACE series performed an earlier FFU than the one of the GERALD series. Bars represent average values ± standard error (SE).

### *SIRPβ1* duplication enhances microglia response in the hippocampus of AD patients

Next, we focused on the impact of the *SIRPβ1* genotype in the patient’s microglia. To address this question, we analyzed the expression of the microglial markers IBA1 and TMEM119 in a series of *post-mortem* human hippocampal samples at Braak V-VI (AD patients) compared to aged-matched control individuals (Braak II without neurological complains) (Fig. 2a and 2b). The results showed that the *SIRPβ1* duplication correlated with an increased area occupied by IBA1 and TMEM119-possitive cells (Mann Whitney U-test, p-value = 0.03) (Fig. 2d and 2e), independent of the amyloid content. Confocal imaging revealed that AD brains (Braak V-VI) bearing the duplication allele showed an increased microglial response (Fig. 2c). Their amyloid plaques were found to be surrounded by more Iba1^+^ cells (Fig. 2f). With regard to the microglial expression signature, patients with the duplication allele exhibited approximately three-times higher levels of *TREM2* (Fig. 2g), which correlates with an increased microglial response. When both activation *(CD45, CSF1, CD74, PU*.*1)* and homeostatic (*CX3CR1, P2RY12, TMEM119*) gene set scores were computed, we observed that the *SIRPβ1* duplication increased both the degree of microglial activation (Fig. 2h) and homeostasis (Fig. 2i) after adjusting by ApoE genotype and Braak stage (Extended Data Table 4).

**Figure 2.**
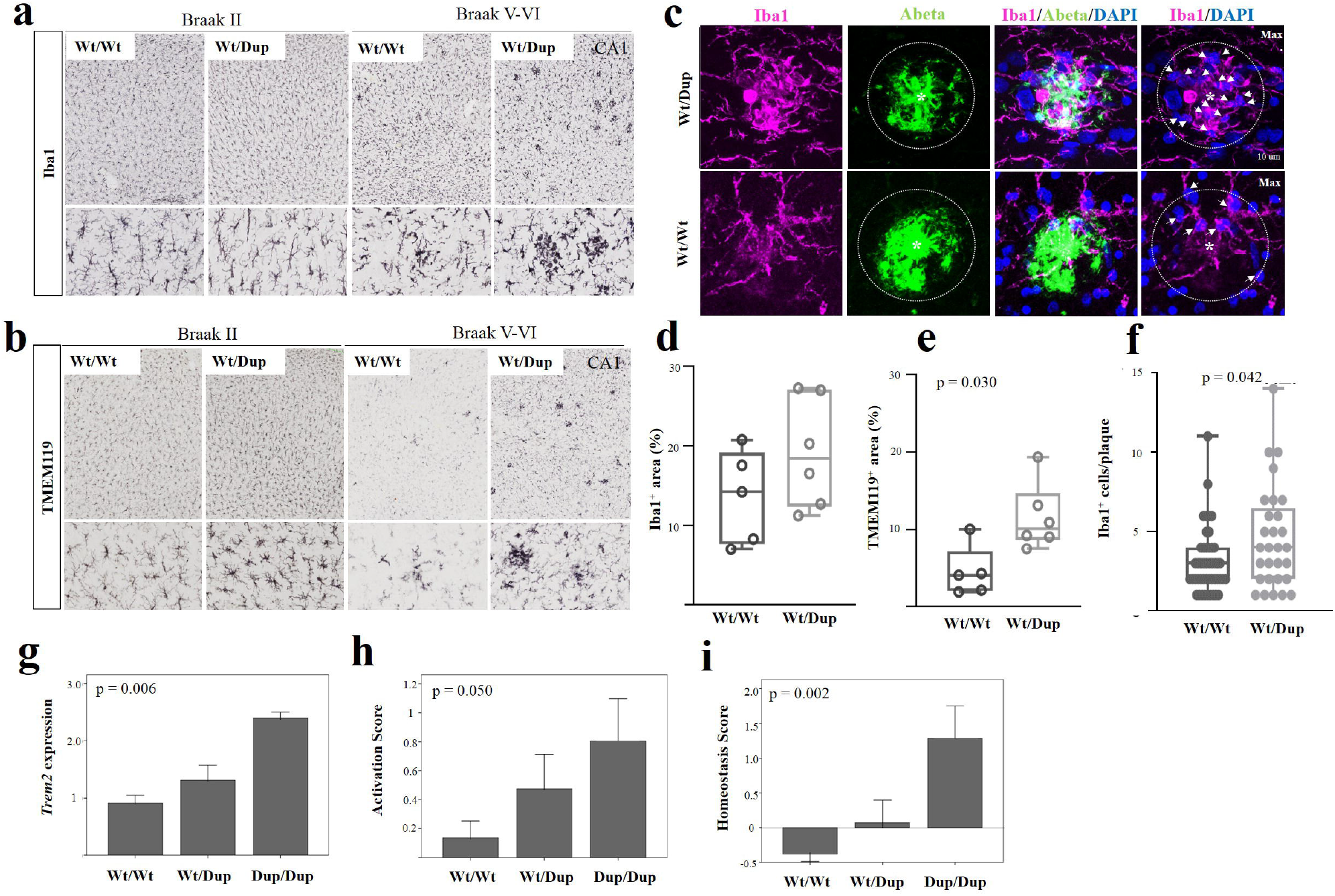
*SIRPβ1* duplication affects plaque associated microglia in AD brains. Effect of SIRPβ1 variant on hippocampal microglia of AD patients. **a**, IBA1 and **b**, TMEM119 immunostaining of illustrative Wt/Wt and Wt/Dup from both Braak II and Braak V-VI subjects. **c**, confocal imaging of three Braak V-VI wild type patients and three Wt/Dup Braak V-VI subjects. A total of 29 and 47 plaques were analyzed respectively. Box plots in **d** and **e** show the IBA1^+^ and TMEM119^+^ area depending on the *SIRPβ1* genotype. **f**, number of IBA1^+^ cells per plaque depending on the *SIRPβ1* genotype. **g**, different average expression (± SE) of *TREM2*. **h** and **i** represent the average gene set scores (± SE) estimated for microglial activation and homeostasis, respectively

### *SIRPβ1* duplication allele changes the isoform landscape altering the Aβ_42_ phagocytosis ability *via* TYROBP

To further examine the impact of the duplication allele on the SIRPβ1 protein structure, we characterized the exact CNV breakpoints. We analyzed a collection of unrelated subjects from the 1,000-genome project with different genetic backgrounds and low-coverage whole-genome sequencing data. We stratified the subjects according to their rs2209313 genotype and represented the coverage according to the hg38 reference genome. We observed a general homogeneity in the breakpoint location regardless of the population origin. Unfortunately, sequencing data did not allow us to clearly define the breakpoints at a nucleotide level (Extended Data Fig. 3). Thus, we analyzed the full sequence of bacterial artificial chromosomes (BAC) which spanned along the critical region. We observed that RP11-77C3 (Genebank AL138804.17) spanned along the entire *SIRPβ1 locus*, including rs2209313, and contained the ancestral haplotype (rs2209313 allele C-CNV Wt allele). In contrast, genome browser versions contain the complete form of the human genome, hence, containing the duplication allele. A local alignment allowed us to define the exact breakpoints of the duplication (Extended Data Fig 3e), and discovered that the structural variant involved exons 2-to-7.

We then explored GTEx RNA-seq data to determine the impact of the duplication allele on the different SIRPβ1 isoforms. *SIRPβ1* is expressed in blood neutrophils, eosinophils and monocytes approximately 1,000-fold higher than in the microglia. Thus, whole-blood GTEx samples were analyzed. According to the ENSEMBL database, there are 12 different protein-coding isoforms. We focused on the four isoforms that were not annotated as “CDS 5’ incomplete” and had different coding sequences. As expected, the presence of rs2209313 allele T-duplication was found to modify *SIRPβ1* isoform spectrum (Fig. 3a). Upon stratification by rs2209313 genotype, we found that the expression of ENST00000381605 (hereafter isoform 205) remained invariable regardless of the donor genotype. However, ENST00000262929 (isoform 201) and ENST00000279477 (isoform 202) were expressed preferentially when the duplication allele was present. On the contrary, ENST00000381603 (isoform 204) disappears with the duplication allele. The detailed analysis of the coding regions revealed that isoforms 201 and 204 contain a single extracellular Ig-like domain, whereas isoforms 202 and 205 display the canonical structure based on three Ig-like domains (Fig. 3b and 3c). Besides, we found two C-terminal regions which affected the TMD and therefore, their potential ability to contact with TYROBP. The amino acid sequence of the isoform 202 TMD (PLL**I**A**F**LLGPK**V**LLVVGVS**V**IY**VY**) is slightly different from that of Isoforms 201, 204 and 205 (PLL**V**A**L**LLGPK**L**LLVVGVS**A**IY**IC**). The major differences are the change of L375 to F375, which does not alter the overall hydrophobic character but introduces a larger aromatic side chain in that position, and the change of C393 to Y393, that actually affects the polarity of the intracellular end of the TMD.

**Figure 3.**
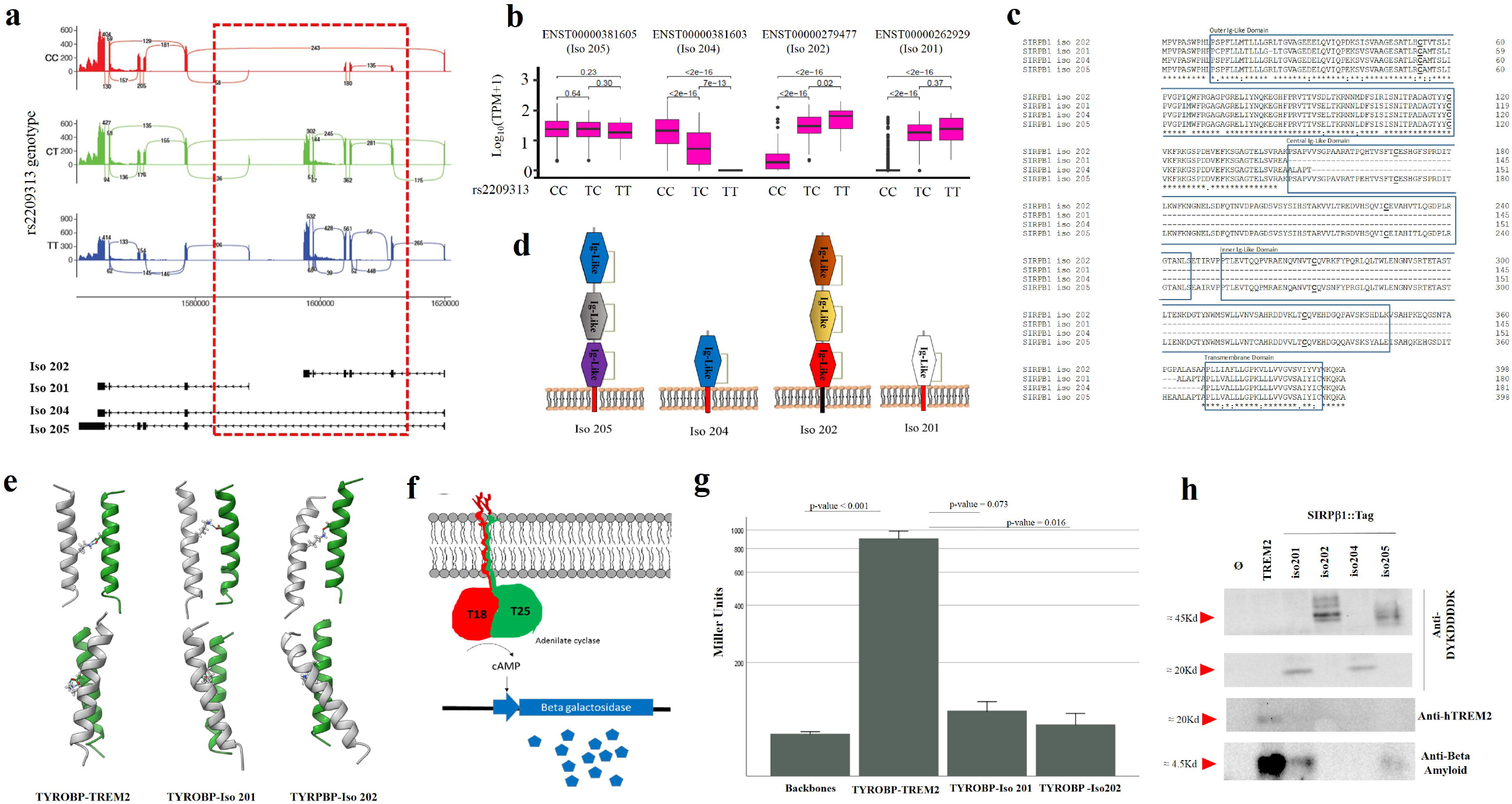
Impact of *SIRPβ1* duplication. Impact of *SIRPβ1* duplication. Panel **a**, Sashimi plot illustrating the Blood-derived RNAseq data available from GTEX according to the rs2209313 genotype. The impact of the duplication can be observed inside the critical region (in dashed red) over each SIRPβ1 isoform. Panel **b**, box plot representation of the relative mRNA abundance of each isoform in blood samples of healthy GTEX donors, according to the *SIRPβ1* rs2209313 genotype. Panel **c** shows the comparison of the four different isoforms that are represented in panel **d**. Panel **e** shows the molecular dynamics results of analyzing TREM2 and SIRPβ1 isoforms 201 and 202 transmembrane domains in the presence of TYROBP’s. Panel **f** illustrates the B2H system used to quantify the transmembrane affinities. Panel **g** shows the Miller Units obtained after the beta-galactosidase assay of liquid cultures co-expressing the empty backbones (n = 4), and TYROBP with TREM2 (n = 6), SIRPβ1 Isoform 201 (n = 6) and SIRPβ1 isoform 202 (n = 6) TMDs. Bars represent the average Miller Units ± SE. Panel **h** sows an illustrative Western Blot of a Aβ_42_ phagocytosis assay of HEK293T cells co-transfected with different plasmid combinations. Lower line corresponds to the intracellular Aβ_42_ while the upper lines correspond to either TREM2 or the different tagged SIRPβ1 isoforms transfected.

In order to explore the structural interaction of the TMD of SIRPβ1 isoforms and TYROBP, we have run Molecular Dynamics (MD) simulations of each dimer in a 1,2-palmitoyl-oleoyl-sn-glycero-3-phosphocholine (POPC) bilayer. We also included TREM2 TMD in the analysis to allow potential comparisons. These TMDs are mainly hydrophobic with a single charged amino acid located in the middle region of the helical structure (K186 and K380 in TREM2 and SIRPβ1, respectively, and D50 in TYROBP) (Fig. 3e, Extended Data Videos 1-3) that interact to create a stable hydrogen bond in all dimers. The orientation of the two domains is however different because of the presence of P379 in the SIRPβ1 isoforms, which introduces a kink right above K380. Also, the positions of K380 in SIRPβ1 and K186 in TREM2 are at slightly different depth into the bilayer, being K186 closer to the extracellular end of the helix, whereas K380 and D50 in TYROBP are at a similar membrane depth. In order to adopt an appropriate angle and distance to form the hydrogen bond between K380 and D50, the helices of SIRPβ1 in the dimer reorient from an almost parallel conformation to a tilted one. This suggests an uneven interaction of the TYROBP-SIRPβ1 depending on the TMD sequence. It is important to point out that in our simulations, SIRPβ1 lacks the large extracellular domain that may potentially restrict the conformation and orientation of the TMD within the lipid bilayer. Taking this into account, we observe a more stable interaction of the TYROBP-SIRPβ1 TMD dimer of isoforms 201, 204 and 205, where the TMD helical axis remains mostly parallel to the membrane normal throughout the simulation, than with the TMD of isoform 202, where the largely tilted orientation may not be reachable in the presence of the extracellular domain.

In order to explore this hypothesis, we cloned the different TMDs together with the membrane-anchoring pf3 N-terminal tag in-frame with the Adenylate Cyclase T18 and T25 domains to perform a membrane-anchored two hybrid analysis (B2H) (Fig. 3f). A first qualitative B2H examination revealed that the TREM2 TMD was more efficient in binding TYROBP than SIRPβ1 TMD. This was evidenced by a more intense X-gal staining of the overnight cultures containing both plasmids (Extended Fig. 4). For a more exhaustive characterization, we performed induced liquid culture β-galactosidase assays, which exhibit a wider dynamic range than the semiquantitative B2H. We observed that the transmembrane interaction between TYROBP and TREM2 TMDs is maximal, with an average of 903 Miller Units (MU), in contrast to the negative control (84 MU). TMD from SIRPβ1 isoform 201 (identical to 204’s and 205’s) was less efficient to contact TYROBP’s TMD (111 MU). As expected by the MD assays, TMD from isoform 202 was highly inefficient in contacting TYROBP’s TMD (94 MU), almost indistinguishable from the negative control (Fig. 3g).

Next, SIRPβ1 flag-tagged isoforms were co-transfected together with TYROBP in HEK293T culture cells (Extended methods). As a positive control, cells co-transfected with plasmids expressing TREM2 and TYROBP were used. Transfectants were then challenged to oligomeric Aβ_42_ in order to evaluate their phagocytosis capacity. Two hours after the challenge, intracellular Aβ_42_ content was determined (Fig. 3h). As expected, HEK293T cell line co-expressing TREM2 and TYROBP gained the ability to phagocyte Aβ_42_ oligomers. SIRPβ1 isoforms 201 and 205 also allowed the HEK293T cells to phagocyte oligomeric Aβ_42_. However, SIRPβ1 isoforms 202 and 204 did not. These data first demonstrate that Aβ_42_ is a ligand of SIRPβ1, although in a clear isoform-dependent manner.

## Discussion

In AD, microglial activation state is either beneficial by phagocytosing Aβ deposits (early disease stages) or harmful as the disease progresses by secreting pro-inflammatory mediators^19^. A defective protective microglial response plays an important role in AD development and progression. Recent transcriptomics analyses have targeted TREM2/TYROBP signaling as the principal regulator of the transformation leading microglia from a homeostatic to a disease-associated state. Furthermore, animal model studies have revealed critical roles for TREM2/TYROBP in the regulation of microglial activity, including survival, phagocytosis, and cytokine production^10,12,14,23,24,25,26^. Previous studies reported SIRPβ1 as a co-receptor of TYROBP, however its role in AD has not been yet fully studied. We have found a common structural variant that alters SIRPβ1 isoform spectrum. The duplication allele precludes the formation of SIRPβ1 isoform 204, while leading to the simultaneous appearance of isoforms 201 and 202. The practical consequence of these changes largely depends on the disease stage. Although meta-analysis shows that the duplication is not a risk factor for AD, MCI patients bearing the Dup/Dup genotype have an increased risk of developing AD as evidenced epidemiologically and determined by their CSF markers. From a structural point of view, brain MRI data show that AD patients carrying the *SIRPβ1* duplication exhibit lower WMH and medial temporal atrophy at the moment of diagnosis, suggesting a worse cognitive response upon the similar neural damage. However, once the disease evolves the effect of the duplication seems to twist. In two independent series, subjects harboring the *SIRPβ1* duplication display a slower cognitive decline. We should highlight that the intimate molecular mechanism underlying this observation remains elusive. We have shown *in vitro* that SIRPβ1 isoforms 201 and 205 have the ability to induce Aβ phagocytosis *via* TYROBP. However, isoforms 202 and 204 ligand-s remain uncharacterized. We could speculate that when the duplication allele is present, the increase of SIRPβ1 isoform 202 together with the suspension of isoform 204 first induces a decrease in the TREM2/TYROBP signaling. This probably reduces microglial phagocytosis in the early stage of the disease. On the contrary, this effect on the TREM2/TYROBP signaling performs important protective functions in the later stages of the disease, maybe limiting local inflammation^27^.

Microglial expression signature of the patients with the *SIRPβ1* duplication shows an increase in homeostatic score, IBA1 and TMEM119-load, probably reflecting an increase in the number of microglial cells, as well as in the microglial activation score. Dup/Dup patients showed nearly three times higher *TREM2* levels than the Wt/Wt. Since TREM2 signaling is involved in microglial activation, metabolic fitness and Aβ plaque protection^28,29^, the increase in *TREM2* levels in Dup patients is consistent with the observed increase in the number of IBA1 positive cells per plaque.

Although overall TREM2/TYROBP-dependent cellular activation appears to be beneficial, the hypothesis of a dual effect depending on the disease phase is not new. Jay et al. suggested the possibility that the functional consequence of TREM2-TYROBP signaling depends on the stage of AD, with detrimental effects in the early stage and beneficial effects in the later ones^30,31^. TREM2-TYROBP is definitely involved in phagocytic clearance, possibly differently at several stages of the process, but the mechanisms remain poorly understood and may depend on the activating ligand^32^. Here we have demonstrated that Aβ_42_ is a ligand of two of the SIRPβ1 isoforms but its role in the *in vivo* interaction between SIRPβ1 and TREM2/TYROBP remains elusive. Recently, Aβ oligomers were reported to bind directly to TREM2 which results in microglial activation and increased expression of proinflammatory cytokines^10^. Neuroinflammation caused by activated microglia was assumed to be detrimental in AD pathology^33^. Our results obtained from patients suffering from mild to moderate cognitive decline suggest the possibility that SIRPβ1-mediated signal could decrease activated microglial toxicity at the early stage of AD, thus strengthening the hypothesis that the association between SIRPβ1 and TREM2/TYROBP counteracts the progressive pathology in AD. This would be in agreement with other studies showing that microglial SIRPβ1 is up-regulated in AD patients and aged APP/J20 transgenic mice, and it is involved in uptake of Aβ_42_. Thus, upregulation of SIRPβ1 isoforms 202 and 201 throughout the course of AD might be a novel target for phagocytic clearance of amyloid plaques^9^.

We are aware of the limitations of our study. First, the cognitive decline has been measured using MMSE taking advantage of its spread use among the different institutions collaborating in the recruitment. We know the limitations of this test and additional longitudinal series with both MMSE and other cognitive indicators must be tested to corroborate our findings. Finally, we should state that the exact molecular mechanisms behind the role of each SIRPβ1 isoform in the pathophysiology of AD will require further investigation.

## Methods

### GERALD

We studied 196 patients with Alzheimer-type dementia attending the Memory Clinic of the Andalusian Institute for Neuroscience in Málaga, Spain. All patients had to meet the criteria of AD according to the benchmarks set forth by the National Institute on Aging Alzheimer’s Association workgroups ^34^ Patients were clinically defined in stages 4, 5 and 6 of the Global Deterioration Scale^35^. Patients who had any evidence suggestive of vascular dementia (VaD), such as focal neurological signs, abrupt deterioration or stepwise progression of cognitive deficits; those with focal vascular lesions (such as hematomas), strokes, normal pressure hydrocephalus; significant neurologic antecedents, such as brain trauma, brain tumors, epilepsy or inflammatory disease; those with serious systemic diseases such as hypothyroidism or chronic renal failure; those with serious psychiatric disorders, substance abuse or developmental anomalies; those who had severe behavioral or communicative issues which would make clinical or MRI examination difficult or those without a reliable informant, were excluded from the study. To avoid the possibility of misclassification of the diagnosis between AD and VaD, we excluded any patient who met the standard criteria for the diagnosis of VaD developed by the National Institute of Neurological Disorders and Stroke and the Association Internationale pour la Recherche et l’Enseignement en Neurosciences. All participants underwent APOE genotyping. Information on demographic and clinical characteristics was collected basally: gender, age, educational level, AD diagnosis, personal and family history, personal psychiatric history, general medication, and antidementia drugs. Cognitive functioning was assessed using the Folstein Mini-Mental State Examination. Cognitive progression was assessed by calculating the slope between the MMSE score obtained at recruitment and the first follow-up.

### GR@ACE

To conduct this research, we recruited 4,656 Spanish individuals; specifically 1,140 sporadic AD patients diagnosed by Neurologists as possible or probable AD in accordance with NINCDS-ADRDA criteria^34^, 1,209 controls with unknown cognitive status from the general population, and 121 neuropsychologically healthy elderly controls who were screened for the absence of cognitive impairment using a structured interview including the neurological mental status examination, category fluency test, and MMSE. Epidemiological data of an initial subset of these series, DNA extraction, and APOE genotyping procedures have been previously described^36^. Ethnicity was Caucasian in AD patients as well as in general population controls. Those individuals with age at onset below 65 years of age were considered as early-onset AD cases.

### rs2209313 Genotyping

Genome-wide association studies meta-analysis on rs2209313 were obtained from the summary file from plink, obtained from different massive genotyping platforms. rs2209313 data from GR@ACE series was extracted from a previous GWAS using the Axiom 815K Spanish Biobank Array^37^, the GERALD series was obtained using specific Taqman probe (Cat. Number 4351379, ThermoFisher Scientific, Massachusetts, USA).

### Brain magnetic resonance imaging

MRI data were acquired on a 1.5 or 3.0 Tesla General Electric Signal scanner. Images were rated by an independent experienced neurologist who was blinded to the clinical data, including cognitive test results and NPI scores. White matter hyperintensities (WMH) and medial temporal atrophy (MTA) were assessed at the moment of diagnosis. WMH and MTA were evaluated as described below. Fluid attenuated inversion recovery and T1-weighted imaging were performed. The extent of WMH severity was rated visually on axial fluid-attenuated inversion recovery images using the Scheltens^38, 39^ visual rating scale for WMH. Frontal periventricular hyper-intensities, occipital periventricular hyper-intensities and lateral periventricular hyper-intensities were each rated on a scale of 0 (no hyper-intensities) to 2 (severe and profuse hyper-intensities), totaling up to a highest possible total score (TPVH) score of 6. In the same way, frontal deep subcortical hyper-intensities, parietal deep subcortical hyper-intensities, temporal deep subcortical hyper-intensities and occipital deep subcortical hyper-intensities were each rated on a scale of 0 (no hyper-intensities) to 6 (severe and profuse hyper-intensities), totaling up to a highest possible total deep subcortical hyper-intensities score of 24. Changes in the basal ganglia, which consist of the caudate, putamen, globus pallidus, thalamus, internal capsule, were considered as white matter lesions and were rated on a scale of 0 (no hyper-intensities) to 30 (severe hyper-intensities). Infratentorial lesions, which consist of cerebellum, midbrain, pons and medulla oblongata, were rated on a scale of 0 (no hyper-intensities) to 24 (severe hyper-intensities). The total WMH score for each patient was the sum of the four scaled ratings. MTA was measured using the Scheltens visual rating scale for MTA.20 The grade of MTA severity was rated visually on coronal T1-weighted and or fluid-attenuated inversion recovery images. Left MTA and right MTA volumes were each rated on a scale of 0 (no atrophy) to 4 (severe atrophy). The total MTA score for each patient was the sum of the two scaled ratings.

### Statistical analysis

For Hardy-Weinberg equilibrium analysis and case control studies, the Institute of Human Genetics (https://ihg.helmholtz-muenchen.de/ihg/snps.html) were used. All polymorphisms included in downstream analysis showed call rates >0.95 and Hardy-Weinberg equilibrium (HWE) (p-value > 0.05). For quantitative analysis IBM SPSS version 25 was used (IBM Corporation, Armonk, New York, USA). Non-normally distributed data were represented using scatter-plots with the median and interquartile range (GraphPad Prism 8.0.1). Two group data were compared by Mann-Whitney U-test. The significance was set at 95% of confidence. To explore association between SIRPβ1 and brain structural imaging measurements we conducted two analysis of covariance (ANCOVA) with MTA and WMH scores as dependent variables. In case of multiple comparisons, Bonferroni’s correction was used. To adequately control their potential confounding role we controlled for HTA and ApoE. A stepwise binary logistic regression analysis, adjusting for MMSE score at diagnosis and ApoE, was fitted to assess associations of SIRPβ1 with cognitive progression. For this purpose, the MMSE slope score was subdivided by median score into those with high or low cognitive decline slope. The Hosmer and Lemeshow’s goodness-of-fit test was applied.

### Alternative splicing analysis

We obtained transcript expression (transcripts per million, TPM) of SIRPβ1 (chr20:1,561,385-1,620,061, reverse strand) in the hippocampus (n = 165) and whole blood (n = 670) from the V8 release of the Genotype-Tissue Expression (GTEx) Project (dbGaP accession phs000424.v8.p2)1. In GTEx, RNA-seq reads are aligned to the human reference genome (build hg38/GRCh38) with STAR2 v2.5.3a, based on the GENCODE v26 annotation. Transcript-level quantifications are obtained with RSEM3 v1.3.0. We also retrieved the genotype at rs2209313 (chr20:1618496, C/T) in the same donors. We restricted our analysis to 4 protein coding transcripts with complete coding sequence (without evidence of 3’ or 5’ end truncation, according to Ensembl): ENST00000279477 (ENST00000568365 has identical CDS), ENST00000381605, ENST00000381603, and ENST00000262929. We assessed the difference in mean transcript expression between genotypes at rs2209313 using the two-sided Wilcoxon Rank-Sum test. We employed Benjamini-Hochberg false discovery rate (FDR) for multiple testing correction, setting the significance level at FDR = 0.05. We represented the distribution of transcript expression (log_10_(TPM+1) for each tissue, transcript, and genotype group at rs2209313 as boxplots. Sample sizes for each tissue and genotype group are the following: blood CC (n = 426), CT (n = 215), TT (n = 29). In boxplots, the box represents the first to third quartiles and the median, and the whiskers indicate ± 1.5 × interquartile range (IQR). All the analyses were performed using R v4.0.3.

### Immunohistochemistry analyses

Human hippocampal autopsy specimens were obtained from the Neurological Tissue Bank of IDIBAPS-Hospital Clinic (Barcelona, Spain). The utilization of post-mortem human samples was approved by the biobank and local ethic committees following Spanish legislation. Subjects were selected on the basis of post-mortem neuropathological examination and clinical data, and all cases were scored for Braak tau pathology. Samples (Extended Data Table 5) included cognitively normal subjects and demented AD patients. For morphological studies, hippocampal regions were fixed in 4% paraformaldehyde in 0.1 M phosphate buffer (PB) for 24-48h, cryoprotected in sucrose, stored at -80ºC, sectioned (30 µm thickness) on a freezing microtome and serially collected in 0.1 M phosphate buffer saline (PBS) and 0.02% sodium azide. Free-floating sections were first heated for general antigen retrieval (80ºC for 20 min in 50 mM citrate buffer at pH 6.0). Then, endogenous peroxidase was inhibited (3% hydrogen peroxide/ 3% methanol in PBS, pH 7.4 for 20 min) and nonspecific staining was avoided by treating with 5% goat serum (Sigma-Aldrich) in PBS. Sections were incubated with the primary antibody over 24-72 hours at room temperature. The antibodies used in this study were: anti-Iba1 rabbit polyclonal (1:1000 dilution, FUJIFILM Wako Pure Chemical Corporation), and anti-TMEM119 rabbit polyclonal (1:500 dilution, Sigma-Aldrich). Tissue-bound primary antibody was then detected by incubating with the corresponding biotinylated secondary antibody (1:500 dilution, 70 min, Vector Laboratories) and streptavidin-conjugated horseradish peroxidase (1:2000, 90 min, Sigma-Aldrich). The peroxidase reaction was visualized with 0.05% 3-3-diaminobenzidine tetrahydrochloride (DAB, Sigma-Aldrich) and 0.01% hydrogen peroxide in PBS. Sections were mounted on SuperFrost Plus™ Adhesion slides (Thermo Scientific), air dried, dehydrated in graded ethanol, cleared in xylene and coverslipped with DPX mounting medium (BDH Laboratories). Sections from control and diseased brains were assayed simultaneously using the same batches of solutions to minimize variability in immunolabeling conditions and the specificity of the immune reactions was controlled by omitting the primary antisera.

For double immunofluorescence staining, sections were incubated sequentially with anti-IBA1 goat polyclonal (1:2000 dilution, Abcam) and anti-amyloid fibrils (OC, 1:5000 dilution, Merck Millipore) followed by the corresponding Alexa 488/568 secondary antibodies (1:1000 dilution, Invitrogen) for 1 hour at room temperature. Subsequent nuclear staining (DAPI dye, Sigma-Aldrich) was performed, and sections were mounted on SuperFrost Plus™ Adhesion slides and coverslipped with 0.01M PBS containing 50% of glycerin and 3% of triethylenediamine (DABCO, Sigma-Aldrich). Treatment with an autofluorescence eliminator reagent (Merck Millipore) following the manufacturer’s recommendations was also performed. Immunofluorescent sections were examined under a confocal laser microscope (Leica SP8). Images are presented as maximum intensity projection of a z-stack with 1 µm steps.

Microglial loading was defined as the percentage of positive area stained with Iba1 (total myeloid cells) or TMEM119 (endogenous microglia) in relation to the total area of the analyzed region. Virtual images of Iba1 or TMEM119 immunostained sections were acquired using the fully automated digital microscopy system dotSlide (Olympus VS120) connected to an Olympus BX61VS optic microscope coupled with a high-resolution digital color camera (VC50 Olympus). The virtual images of the whole tissue sections were digitized at 40x magnification producing virtual images with a resolution of 0.173 μm/pixel and an average image size of 34213 pixel x 46480 pixel. Images from Braak V-VI hippocampus (including dentate gyrus and CA1-4) were then acquired using the Olyvia 2.6 image viewer software (Olympus) (image size: 1654 pixel x 877 pixel; pixel size: 28.34 pixel/cm). Digital images (2 sections/individual, n=5-6) were processed using the Visilog 6.3 (Noesis) image analysis system. The Iba1-or TMEM119-immunopositive signal within the selected brain region was converted into 8-bit gray scale, and immunostained microglial cells were identified by a threshold level mask. With the intensity threshold, only pixels with a staining intensity above a certain value in the intensity range scales (0-255) were visualized. A fixed threshold level (ranging between 170-180) was maintained throughout the image analysis of all sections from the same individual brain for uniformity.

To analyze the number of IBA1-positive cells surrounding amyloid plaques, double IBA1/OC immunofluorescence sections stained with DAPI were used. Amyloid plaques from Braak VI hippocampus were randomly selected (29-47 plaques, n=3-4 individuals) and z-stack confocal images were acquired with 63X 1.4 NA objective and 1 µm steps. All optical sections of the z-stack were quantified and plaque-associated Iba1 cells (including a 20µm-extra radius from the plaque edge) were counted.

### Total RNA extraction

Total RNA were sequentially extracted, from human hippocampal tissue, using TRIsure Isolation Reagent (Bioline) (100□mg tissue/1□ml TRIsure Isolation Reagent) and purified by RNasy Mini Kit (Qiagen) following manufacture recommendations^40^. RNA integrity was determined by RNA Nano 6000 (Agilent) (RIN: 4.95 ± 1.4). RNA was quantified using Nanodrop 2000 (Thermo Fischer).

### Retrotranscription and quantitative real-time RT-PCR

Reverse transcription (RT) (4□μg of total RNA as template) was performed with High Capacity cDNA Archive Kit (Applied Biosystems). For real time qPCR, 40□ng of cDNA were mixed with 2× Taqman Universal Master Mix (Applied Biosystems) and 20× Taqman Gene Expression assay probes (Applied Biosystems). Quantitative RT-PCR (qPCR) was done using Taqman probes (Applied Biosystems). The different cDNAs were mixed with Taqman Universal Master Mix (Applied Biosystems) and Taqman Gene Expression assay probes. The quantitative PCR reactions (qPCR) were done using an ABI Prism 7900HT (Applied Biosystems). For activation or homeostatic set score, the expression of CD45 (Hs04189704_m1),d45 CSFsf1 (Hs00174164_m1), CDd74 (Hs00269961_m1) and PUu.1 (Hs02786711_m1) or CX3CRx3cr1 (Hs01922583_s1), P2RYry12 (Hs01881698_m1) and TMEMmem119 (Hs01938722_u1) were determined, respectively. Results were always expressed using the comparative double-delta Ct method where ΔCt values represent GAPDH normalized expression levels. Gene set score was calculated as described^41^.

## Data Availability

All data produced in the present study are available upon reasonable request to the authors upon the approval of the ethic's review board.

## Acknowledgements

The authors would like to thank patients, controls and caregivers their participation in the GERALD and GR@ACE/DEGESCO consortium initiatives. This project did not receive any specific public nor private funding. It has been possible only thanks to the goodwill and the collaborative effort of the researchers and institutions signing this paper. RG acknowledge the support of the Spanish Ministry of Science and Innovation to the EMBL partnership, the Centro de Excelencia Severo Ochoa and the CERCA Programme/Generalitat de Catalunya. RG is principal investigator from grant PGC2018-094017-B-I00 from AEI/FEDER, EU. AG is principal investigator of Instituto de Salud Carlos III (ISCIII) of Spain grants PI18/01557 and P21/00915 co-financed by the FEDER funds from European Union. JV is principal investigator of ISCIII grants PI18/01556 and PI21/00914 co-financed by FEDER funds from European Union, and Junta Andalucia grant US-1262734 co-financed by Programa Operativo FEDER 2014-2020. JJT is principal investigator of PID2019-103921GB-I00 from the Spanish Ministerio de Ciencia e Innovación. JLR is principal investigator of UMA20-FEDERJA-133 from the program FEDER Andalucía 2014-2020, EU. JLR also acknowledges the Centro de Supercomputación y Bioinformática (Picasso-server) for computing support. I. de Rojas is supported by national grant from the Instituto de Salud Carlos III FI20/00215. CM-C was supported by FPU fellowship from the Spanish Ministry of Science, Innovation, and Universities. The Genome Research @ Fundació ACE project (GR@ACE) is supported by Grifols SA, Fundación bancaria “La Caixa”, Fundació ACE, and CIBERNED. A.R. and M.B. receive support from the European Union/EFPIA Innovative Medicines Initiative Joint undertaking ADAPTED and MOPEAD projects (grant numbers 115975 and 115985, respectively). M.B. and A.R. are also supported by national grants PI13/02434, PI16/01861, PI17/01474, PI19/01240 and PI19/01301. Acción Estratégica en Salud is integrated into the Spanish National R□+□D□+□I Plan and funded by ISCIII (Instituto de Salud Carlos III) —Subdirección General de Evaluación and the Fondo Europeo de Desarrollo Regional (FEDER—”Una manera de hacer Europa”). Some control samples and data from patients included in this study were provided in part by the National DNA Bank Carlos III (www.bancoadn.org<http://www.bancoadn.org>, University of Salamanca, Spain) and Hospital Universitario Virgen de Valme (Sevilla, Spain); they were processed following standard operating procedures with the appropriate approval of the Ethical and Scientific Committee. We would like to dedicate this work to the memory of Dr. Jose Luis Gómez-Skarmeta, colleague and friend who always supported the project.

