## Supplementary figures and images for "An intragenic duplication within *SIRPβ1* shows a dual effect over Alzheimer’s disease cognitive decline altering the microglial response"

### Extended Figure 1

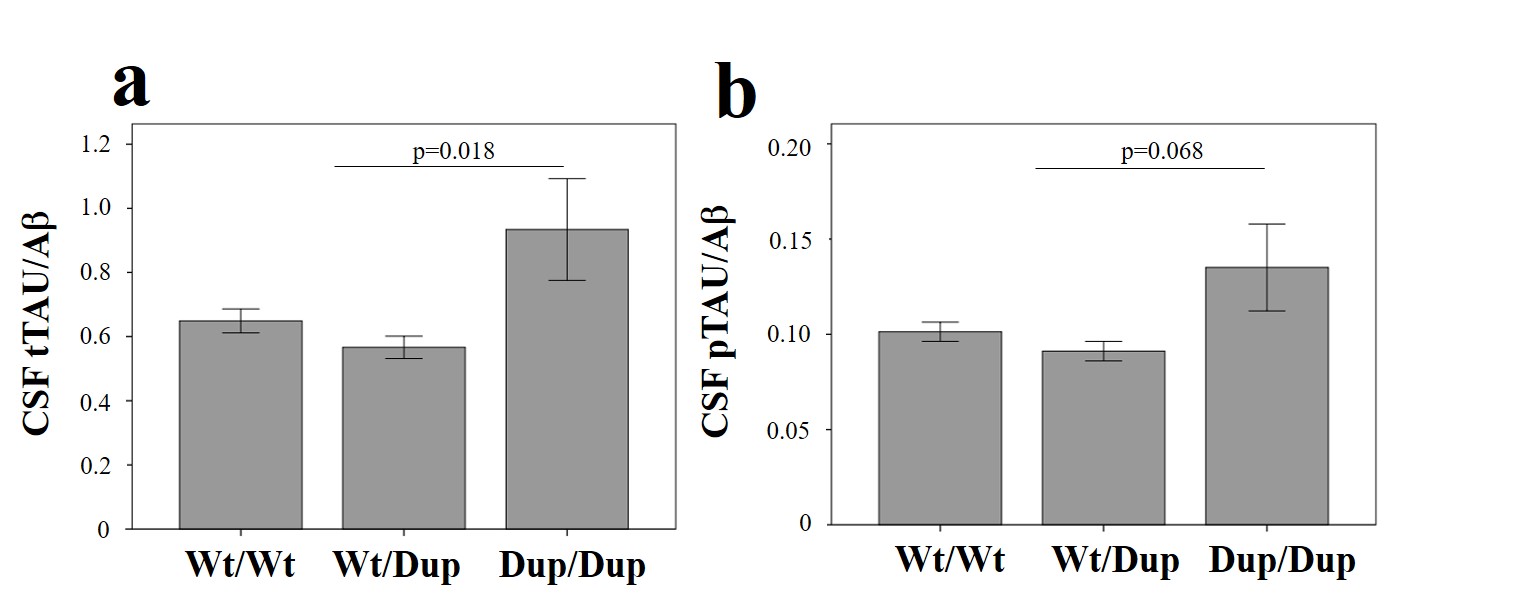

### Extended Figure 2

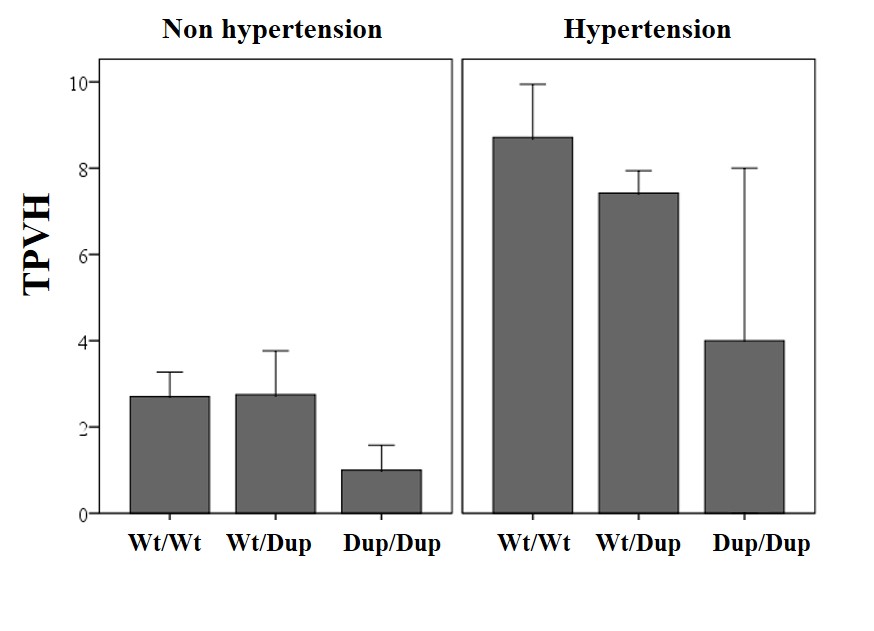

### Extended Figure 3

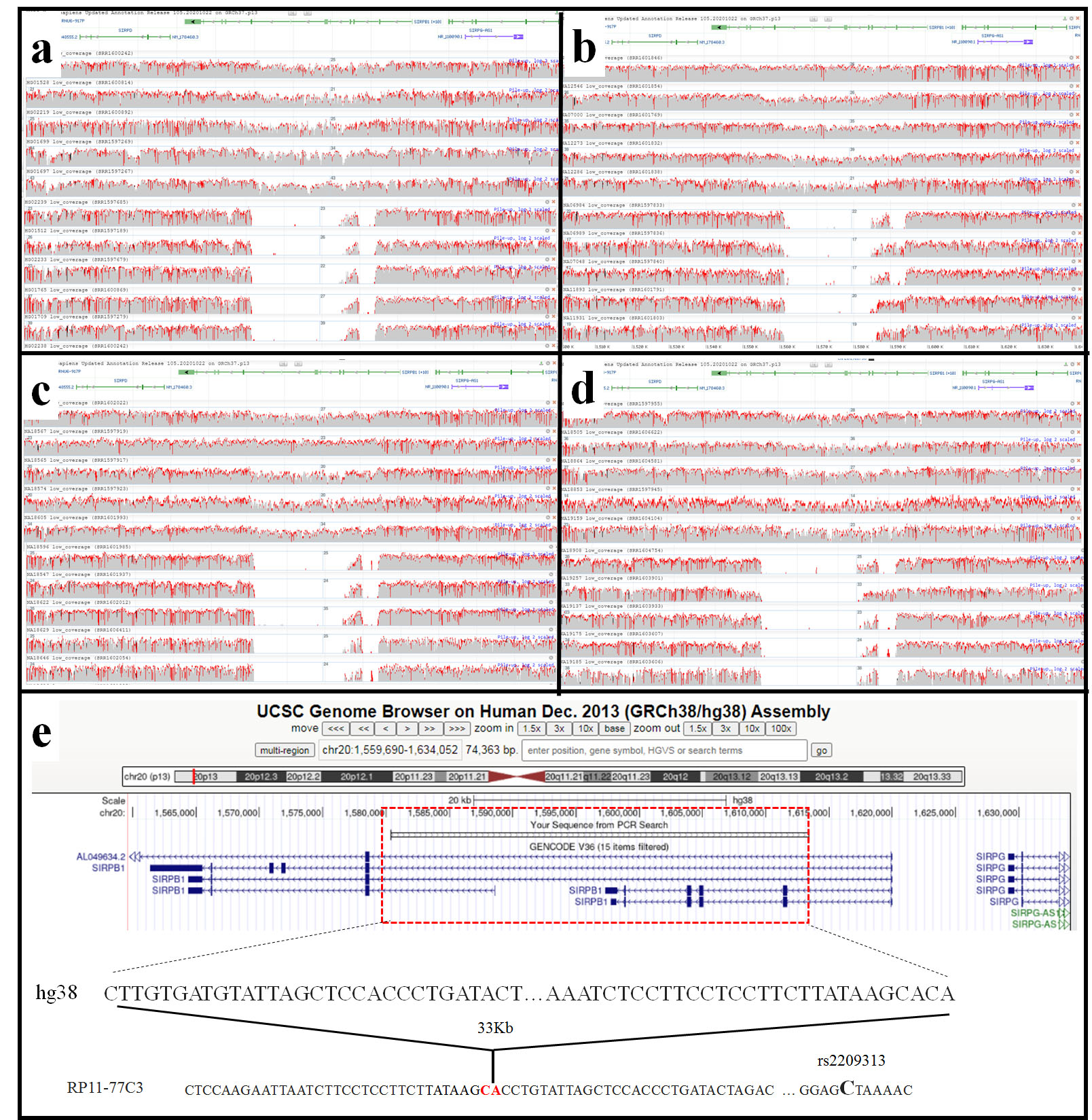

### Extended Figure 4

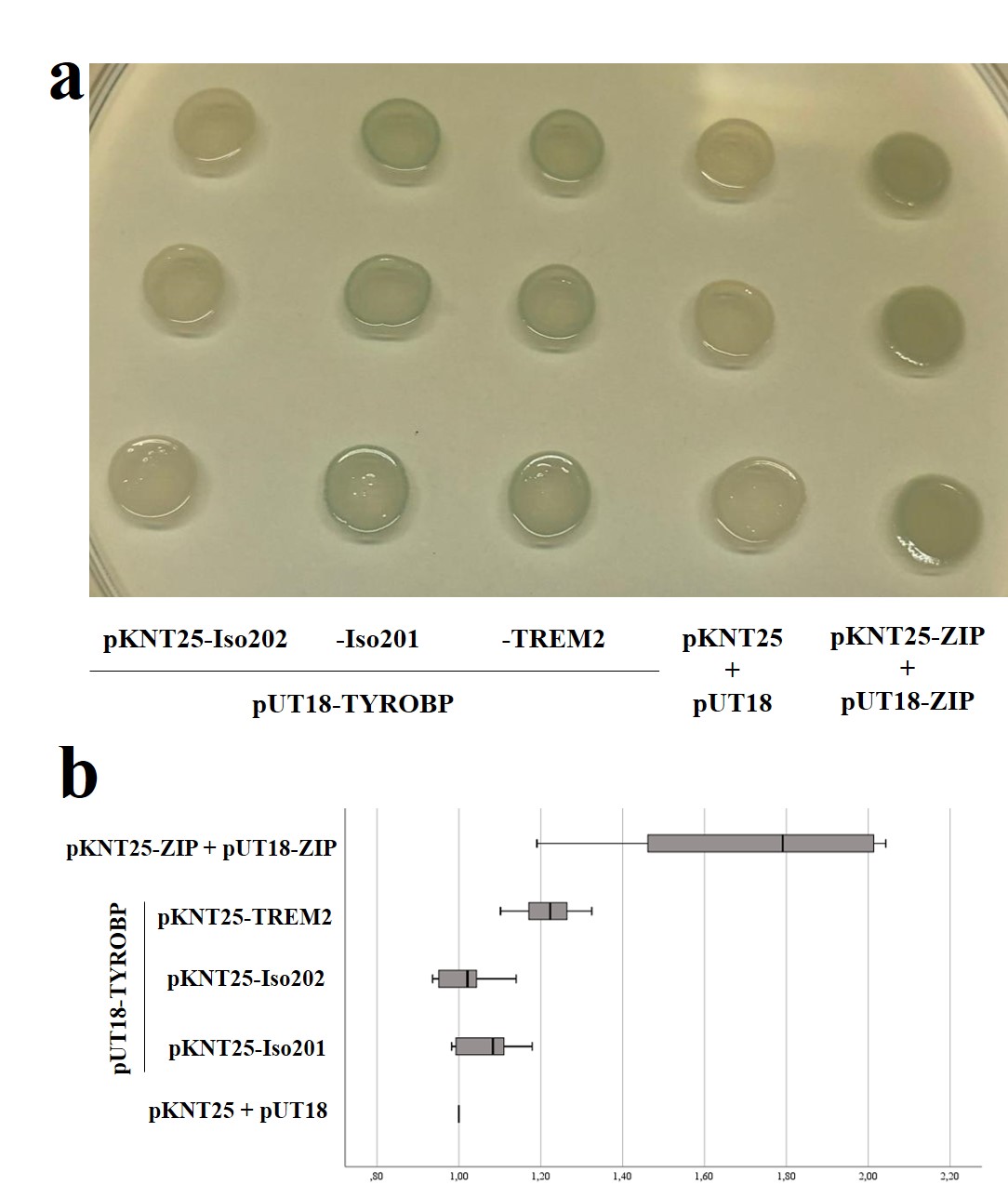
