## Extended data and Methods for "An intragenic duplication within *SIRPβ1* shows a dual effect over Alzheimer’s disease cognitive decline altering the microglial response"

José María García-Alberca et al.

**Extended Data**

**Extended Data Fig. 1. Cerebrospinal fluid analysis of MCI patients according to their *SIRPβ1* genotype.** **a** Total tau/Aβ ratio and **b** Phospho-tau/Aβ ratio of Mild cognitive impairment patients according to the rs2209313 genotype (Wt/Wt, n=307; Wt/Dup=171; Dup/Dup=15). Bars represent the average values ± standard errors (SE). p-values were calculated with Mann Whitney U-test.

**Extended Data Fig. 2. Deep WHM according to the *SIRP****β****1* genotype.** Correlation between *SIRP*β*1* rs2209313 genotype and the hypertensive status of the GERALD AD patients. Bars represent the average values ± standard errors (SE)

**Extended Data Fig. 3. Characterization of the *SIRP***β***1* CNV breakpoints.** Panels **a** to **d** show the sequencing coverage of 40 unrelated donors of the 1,000 genome project from CEU, IBS, HAN and YRI populations, respectively. On each panel the top five subjects show rs2209313 T/- genotype (duplication allele) and the bottom five subjects the rs2209313 CC genotype (Wild type CNV allele). Panel **e** shows the exact breakpoints thanks to the local alignment of the full sequence of the RP11-77C3 BAC and the hg38 genome.

**Extended Data Fig. 4. Semiquantitative B2H.** Panel **a** shows the X-gal induced color of overnight triplicate drops of BTH101 *E. Coli* strain co-expressing both the pUT18 and pKNT25 derivates. As a positive control, both plasmids contained a translational fusion to ZIP, leucine zipper of GCN4 which dimerizes in the cytoplasm. Color intensities were referred to the one showed by the strain expressing the empty backbones. TYROBP, SIRPβ1 and TREM2 transmembrane domains were anchored to the plasma membrane via a pf3 tag. Panel **b** shows a graphical representation of the mean color intensity upon normalization to the empty backbones (n=12). Whiskers show the total range and the boxes the interquartile range. TREM2 transmembrane domain (n=8) was found to be more efficient in its contact with TYROBP transmembrane domain when compared to SIRPβ1’s (n=9 each).

**Extended Data Video 1**. Molecular dynamics showing a representative TYROBP TREM2 TMD interaction in a POPC bilayer

**Extended Data Video 2.** Molecular dynamics showing a representative TYROBP-SIRPβ1 iso202 TMD interaction in a POPC bilayer

**Extended Data Video 3.** Molecular dynamics showing a representative TYROBP-SIRPβ1 iso201 TMD interaction in a POPC bilayer

**Extended Data Table 1. Role of *SIRP***β***1* rs2209313 in AD risk**

| **Study** | **Cohort** | **Size** | **rs2209313-T MAF** | **OR [CI_95_]** | **p-value** |
| --- | --- | --- | --- | --- | --- |
| **Moreno-Grau et al. 2019**^1^ | DEGESCO | 15,894 | 0.200 | 0.958 [0.901-1.015] | 0.140 |
| **Kunkle et al. 2019**^2^ | IGAP | 63,926 | 0.212 | 0.966 [0.931-1.001] | 0.051 |
| **Marioni et al. 2018** | UK biobank | 548955 | N.A. | 0.997 [0.964-1.031] | 0.871 |
| **Furney et al. 2011**^3^ | ADDN1 | 325 | 0.197 | 1.064 [0.596-1.532] | 0.794 |
| **Furney et al. 2011**^3^ | ADDN2 | 312 | 0.202 | 0.987 [0.570-1.403] | 0.950 |
| **Antunez et al. 2011**^4^ | Neocodex | 1,078 | 0.208 | 1.101 [0.715-1.488] | 0.624 |
| **Meta-analysis** | Merged | 630,049 |  | 0.980 [0.960-1.000] | **0.06** |

**Extended Data Table 2. MCI conversion risk on the GR@CE series**

|  | **Dementia** | | **Alzheimer** | |
| --- | --- | --- | --- | --- |
| **Variable** | **OR** | **p-value** | **OR** | **p-value** |
| *SIRPβ1* Dup/Dup | 1.544 | 0.008 | 1.678 | 0.018 |
| Age | 1.093 | <0.001 | 1.093 | <0.001 |
| Sex | 1.061 | 0.413 | 1.413 | <0.001 |
| ApoE4 dose | 1.682 | <0.001 | 2.196 | <0.001 |

**Extended Data Table 3. GERALD and GR@CE series**

| Series |  | *SIRP*β*1*  rs2209313  Genotype (n) | | | ApoE Genotype (n) | |  |  |  |
| --- | --- | --- | --- | --- | --- | --- | --- | --- | --- |
|  |  |  |  |  |  |  | MMSE at Diagnosis Average  [IC_95_] | MMSE Slope  Average  [IC_95_] | FFU  Average months  [IC_95_]) |
|  |  | CC | TC | TT | ApoE4^-^ | ApoE4^+^ |  |  |  |
| GERALD | Male | 23 | 17 | 3 | 25 | 18 | 21.53 [13-29] | -0.12 [-0.50-0.45] | 20.11 [3-59] |
|  | Female | 84 | 48 | 6 | 65 | 70 | 17.15 [3-26] | -0.13 [-0.65-0.50] | 17.63 [4-61] |
| GR@CE | Male | 856 | 392 | 52 | 731 | 569 | 20.29 [10-27] | 0.18 [-0.40-0.87] | 10.08 [5.75-14.98] |
|  | Female | 2,181 | 1,037 | 138 | 1,950 | 1,406 | 19.12 [11-27] | 0.16 [-0.33-0.74] | 10.19 [6.18-15.15] |

**Extended Data Table 4. Transcriptional analysis according to the *SIRP****β****1* genotype**

|  | Entire series | | Braak V-VI | | Braak 0-IV | |
| --- | --- | --- | --- | --- | --- | --- |
| Microglial Activation gene set score | Beta | p-value | Beta | p-value | Beta | p-value |
| *SIRP*β*1* rs2209313 T-Dup allele | **0.232** | **0.043** | **0.374** | **0.05** | 0.115 | 0.539 |
| Stage | **0.493** | **<0.001** | - | - | 0.180 | 0.340 |
| Microglial Homeostasis gene set score | Beta | p-value | Beta | p-value | Beta | p-value |
| *SIRP*β*1* rs2209313 T-Dup allele | **0.362** | **0.005** | **0.561** | **0.002** | 0.191 | 0.311 |
| Stage | -0.104 | 0.405 | - | - | 0.084 | 0.654 |
| Normalized *Trem2* expression | Beta | p-value | Beta | p-value | Beta | p-value |
| *SIRP*β*1* rs2209313 T-Dup allele | **0.336** | **0.009** | **0.537** | **0.006** | 0.136 | 0.464 |
| Stage | 0.127 | 0.315 | - | - | 0.226 | 0.227 |

**Extended Data Table 5. Demographic data for *post-mortem* human brains.**

| **Case** | **Braak stage** | **Age range (years)** | **Gender** | ***Post-mortem* delay (hours)** | ***SIRP****β****1* CNV** | **Neuropathological diagnosis by Biobank** |
| --- | --- | --- | --- | --- | --- | --- |
| Case 1 | II | 81-85 | Female | 7.5 | Wt/Dup | Small vessel disease (SVD). Neurofibrillary pathology |
| Case 2 | II | 76-80 | Female | 9.5 | Wt/Dup | Hepatic encephalopathy. Alzheimer's disease related pathology (ARP) |
| Case 3 | II | 76-80 | Male | 6.5 | Dup/Dup | SVD with infarct. ARP |
| Case 4 | II | 76-80 | Female | 10.5 | Wt/Wt | SVD |
| Case 5 | II | 91-95 | Female | 20 | Wt/Wt | SVD. Neurofibrillary pathology |
| Case 6 | V-VI | 81-85 | Female | 6 | Wt/Dup | Memory disorder. Amyloidopathy. Alpha-synuclein in olfactory bulb. Adenocarcinoma brain metastasis. |
| Case 7 | V-VI | 76-80 | Male | 12.5 | Wt/Dup | Conduct disorder. Amyloidopathy. CAA severe |
| Case 8 | V-VI | 35-36 | Male | 15 | Wt/Dup | Amyloidopathy. CAA. Lewy bodies. Mutation Ile716Phe in APP |
| Case 9 | V-VI | 56-60 | Male | 7 | Wt/Dup | Depression. Amyloidopathy. CAA. Mutation p264L in PS1 |
| Case 10 | V-VI | 81-85 | Male | 8 | Wt/Dup | Memory disorder. Amyloidopathy. Alpha-synuclein. Limbic-predominant age-related TDP-43 encephalopathy. Neocortical Lewy bodies. Hippocampal sclerosis. |
| Case 11 | V-VI | 81-85 | Male | 4 | Wt/Dup | Memory disorder. Amyloidopathy. CAA severe. Lobar hemorrhage. Hepatitis C. |
| Case 12 | V-VI | 81-85 | Male | 3 | Wt/Wt | Memory disorder. Amyloidopathy.  Vascular encephalopathy. SVD with microinfarcts |
| Case 13 | V-VI | 61-65 | Male | 6.5 | Wt/Wt | Memory disorder. Amyloidopathy. CAA severe. Lewy bodies. SVD.  Down´s Syndrome. |
| Case 14 | V-VI | 66-70 | Female | 7 | Wt/Wt | Memory disorder. Amyloidopathy |
| Case 15 | V-VI | 81-85 | Female | 6 | Wt/Wt | Memory disorder. Amyloidopathy. CAA |
| Case 16 | V-VI | 61-65 | Female | 8 | Wt/Wt | Memory disorder. Amyloidopathy. |

**Extended** **Methods**

Jose Maria García-Alberca et al

**Plasmids and strains.** In order to evaluate the affinity of the different transmembrane domains for TYROBP, we carried out two-hybrid assays in bacteria. In order to generate a series of hybrid clones that contain a signal peptide to anchor the transmembrane domain in the plasma membrane, as well as the two domains whose complementation results in an indicator activity. Constructs contained the Pf3 signal peptide (QSVITDVTGQLTAVQADITTIGG) followed by the transmembrane domain of the protein (hereafter named TYROBPTM, TREM2TM, SIRPβ1iso201TM and SIRPB1iso202TM) and a 3-glycine elbow to increase flexibility in order to facilitate the interaction between the two adenylate cyclase domains of the BATCH dual hybrid system. Plasmids containing the chimeric fusions were ordered from Genescript (www. <https://www.genscript.com/>). and further subcloned using conventional molecular biology techniques in DH5α *E. Coli* strain. The complete protein used as bait is translated from plasmid T25 of the in-frame cloned BATCH pKTN25 plasmid, at the BamHI and HindIII sites, generating the full construct Pf3: TYROBPTM:

MTMITPSLQSVITDVTGQLTAVQADITTIGGGVLAGIVMGDLVLTVLIALAVYFLGGGDPRVPSSNSMTMQQSHQAGYANAADRESGIPAAVLDGIKAVAKEKNATLMFRLVNPHSTSLIAEGVATKGLGVHAKSSDWGLQAGYIPVNPNLSKLFGRAPEVIARADNDVNSSLAHGHTAVDLTLSKERLDYLRQAGLVTGMADGVVASNHAGYEQFEFRVKETSDGRYAVQYRRKGGDDFEAVKVIGNAAGIPLTADIDMFAIMPHLSNFRDSARSSVTSGDSVTDYLARTRRAAPSI

The full protein translated as prey from the plasmid pT18 of the BATCH kit upon an equivalent cloning generated the Pf3: TREM2TM construct:

MTMITPSLQSVITDVTGQLTAVQADITTIGGSILLLLACIFLIKILAASALWAGGGDPRVPSSNSAASEATGGLDRERIDLLWKIARAGARSAVGTEARRQFRYDGDMNIGVITDFELEVRNALNRRAHAVGAQDVVQHGTEQNNPFPEADEKIFVVSATGESQMLTRGQLKEYIGQQRGEGYVFYENRAYGVAGKSLFDDGLGAAPGVPSGRSKFSPDVLETVPASPGLRRPSLGAVERQSI

The complete protein that is translated from the plasmid T18 of the BATCH kit cloned with the construct Pf3:SIRPB1iso201TM is:

MTMITPSLQSVITDVTGQLTAVQADITTIGGPLLVALLLGPKLLLVVGVSAIYICGGGDPRVPSSNSAASEATGGLDRERIDLLWKIARAGARSAVGTEARRQFRYDGDMNIGVITDFELEVRNALNRRAHAVGAQDVVQHGTEQNNPFPEADEKIFVVSATGESQMLTRGQLKEYIGQQRGEGYVFYENRAPSSIYGVAGKSLVGAFGAVPSSIGVAGKSLVGAFGAVPSSIGVAGKSLVGAFGAVPSSIGVAGKSLVGAFGAPSSIYGVAGKSL

Finally, the complete protein that is translated from the plasmid T18 of the BATCH kit cloned with the construct Pf3SIRPB1Iso202TM is:

MTMITPSLQSVITDVTGQLTAVQADITTIGGPLLIAFLLGPKVLLVVGVSVIYVYGGGDPRVPSSNSAASEATGGLDRERIDLLWKIARAGARSAVGTEARRQFRYDGDMNIGVITDFELEVRNALNRRAHAVGAQDVVQHGTEQNNPFPEADEKIFVVSATGESQMLTRGQLKEYIGQQRGEGYVFYENRAPSSIYGVAGKSLVGAFGAVPSSIGVAGKSLVGAFGAVPSSIGVAGKSLVGAFGAVPSSIGVAGKSLVGAFGAPSSIYGVAGKSL

**Cell culture transfection and phagocitosis assay.** HEK293T cells (ATCC, Manassas, VA, United States) and maintained in DMEM medium supplemented with 10% fetal bovine serum, GlutaMAX, and 1% penicillin/streptomycin/fungizone (Thermo Fisher Scientific, Waltham, MA, United States). For transfection, HEK293T were grown on 24-well plates until reaching 70–80% confluence and transfected using lipofectamine 2000 (Invitrogen, Carlsbad, CA, United States). Plasmids were designed *in silico* overthe pCDNA3.0 backbone and obtained from Genscript ([www.genscript.com](http://www.genscript.com)). Expression was driven from CMV promoter. SIRPβ1 Isoform 201 sequence was: MPVPASWPHPPCPFLLLTLLLGLTGVAGEDELQVIQPEKSVSVAAGESATLRCAMTSLIPVGPIMWFRGAGAGRELIYNQKEGHFPRVTTVSELTKRNNLDFSISISNITPADAGTYYCVKFRKGSPDDVEFKSGAGTELSVREAALAPTAPLLVALLLGPKLLLVVGVSAIYICWKQKA**DYKDDDD**K. Isoform 202 MPVPASWPHLPSPFLLMTLLLGRLTGVAGEEELQVIQPDKSISVAAGESATLHCTVTSLIPVGPIQWFRGAGPGRELIYNQKEGHFPRVTTVSDLTKRNNMDFSIRISNITPADAGTYYCVKFRKGSPDHVEFKSGAGTELSVRAKPSAPVVSGPAARATPQHTVSFTCESHGFSPRDITLKWFKNGNELSDFQTNVDPAGDSVSYSIHSTAKVVLTREDVHSQVICEVAHVTLQGDPLRGTANLSETIRVPPTLEVTQQPVRAENQVNVTCQVRKFYPQRLQLTWLENGNVSRTETASTLTENKDGTYNWMSWLLVNVSAHRDDVKLTCQVEHDGQPAVSKSHDLKVSAHPKEQGSNTAPGPALASAAPLLIAFLLGPKVLLVVGVSVIYVYWKQKA**DYKDDDDK.** Isoform 204 MPVPASWPHLPSPFLLMTLLLGRLTGVAGEDELQVIQPEKSVSVAAGESATLRCAMTSLIPVGPIMWFRGAGAGRELIYNQKEGHFPRVTTVSELTKRNNLDFSISISNITPADAGTYYCVKFRKGSPDDVEFKSGAGTELSVREAALAPTAPLLVALLLGPKLLLVVGVSAIYICWKQKA**DYKDDDDK** and Isoform 205: MPVPASWPHLPSPFLLMTLLLGRLTGVAGEDELQVIQPEKSVSVAAGESATLRCAMTSLIPVGPIMWFRGAGAGRELIYNQKEGHFPRVTTVSELTKRNNLDFSISISNITPADAGTYYCVKFRKGSPDDVEFKSGAGTELSVRAKPSAPVVSGPAVRATPEHTVSFTCESHGFSPRDITLKWFKNGNELSDFQTNVDPAGDSVSYSIHSTARVVLTRGDVHSQVICEIAHITLQGDPLRGTANLSEAIRVPPTLEVTQQPMRAENQANVTCQVSNFYPRGLQLTWLENGNVSRTETASTLIENKDGTYNWMSWLLVNTCAHRDDVVLTCQVEHDGQQAVSKSYALEISAHQKEHGSDITHEAALAPTAPLLVALLLGPKLLLVVGVSAIYICWKQKA**DYKDDDDK**. To assay the phagocytic capacity, 48 hours post-transfection cells were challenged with a solution of 1 μM oligomeric Aβ. Aβo were prepared as previously described by Sadkleir and coworkers. Briefly, lyophilized recombinant Aβ_42_ peptide (rPeptide, A-1163) was re-suspended to 5 mM in DMSO and then further diluted to 100 μM in in sterile PBS. The peptide was incubated at 4 °C for 24 h to generate oligomers. Cells were incubated dor 2h at 37ºC and washed twice with saline and resuspended in TRIsure^TM^, (Bioline, United Kingdom).

**Western blot analysis.** Proteins were fractionated by electrophoresis using 10% sodium dodecyl sulphate (SDS) polyacrylamide gels, electroblotted into PVDF membranes (Hybond-P, GE Healthcare), and blocked with 5% BCA in TBS. Membranes were then incubated with the different antibodies overnight at 4 °C (anti-hTREM2, anti-dykddddk from Cell Signalling were used according to manufacturer’s instructions), followed by incubation with a horseradish peroxidase-conjugated antibody. The immunoreactive bands were visualized using ECL (Invitrogen). The images were obtained and analyzed with ChemiDoc™ Touch Imaging System (Bio‐Rad). To analyze Aβ, protein samples were loaded onto 16% SDS-Tris-Tricine-PAGE and transferred to PVDF (Inmobilon-P, Millipore). After blocking, using 5% non-fat milk, the membranes were incubated overnight, at 4ºC, with a mixture of N-terminus (clone 82E1) mouse monoclonal (1:1000, IBL) and anti-Aβ (clone 6E10) mouse monoclonal (1:1000, Signet).

**Semiquantitative Bacterial Two Hybrid.** The test was carried out on the BTH101 strain (Str^R^. *lacZ*^+^, F^-^, cya-99, araD139, galE15, galK16, rpsL1, hsdR2, mcrA1, mcrB). Reconstruction of the adenylate cyclase activity was done thanks to the interaction of the tested domains. This increases the levels of cyclic AMP, which is translated into an increase in the expression of the β-galactosidase. TSS-competent BTH101 were prepared and stored at -80 ° C. Prior to each assay, plasmids were freshly transformed in BTH101 and plated at 37 ° C overnight. Four colonies were selected from each plate, and added separately to 1 mL of LB with 100 ng/µL ampicillin and 50 ng/µL of kanamycin, and stirred at 1200 rpm overnight at 30 ° C. The next day, 10 µL drops of each biological replica were seeded in triplicate on plates with both antibiotics and grown at 30°C. After 24h, a picture was taken to analyze the intensity of blue of each drop with the ImageJ software. The blue color will be proportional to the amount of cAMP generated, linked to the interaction capacity of the study proteins, and was normalized with respect to the negative control (BTH101 with pUT18 and pKNT25). For the analysis, a 32-bit image type was selected, the different rows were defined with the Analyze Gel command, then the baseline was defined with Plot Lanes and the interior of the peak area was chosen to obtain the result.

**Quantitative Bacterial Two Hybrid.** Following the same strategy, freshly transformed BTH101 with the necessary plasmids were grown overnight at 37°C. Next, a fresh 1:50 dilution in 1.5 mL of LB with both ampicillin and kanamycin. Dilutions were grown in 10 mL tubes at 30°C and 180 rpm. When OD_600_ reached 0.2-0.3, 1.5 µL of IPTG (20 ng / mL) was added to 0.75 mL of culture and allowed to grow overnight. Next, 100 µL aliquots were taken, diluted 1:10 and OD_600_ was measured. 50 µL of the diluted samples were taken and added to 705 µL of buffer Z (0.06 M Na_2_HPO_4_ · 7H_2_O; 0.04 M NaH_2_PO4 · H_2_O; 0.01 M KCl; 0.001MgSO_4_ 7H_2_O) with ß-mercaptoethanol (135 µL of ß-mercaptoethanol were added to each 50 mL of buffer Z), 30 µL of chloroform and 15 µL of 0.1% SDS. As a negative control, sterile LB was used instead of saturated culture. Samples were vortexed for 10 s, and stabilized for 2 min in a water bath at 30°C. The reaction was started by adding 200 µL ONPG (4 mg / mL in buffer Z), and the time it took to turn yellow was recorded (t), at which point it was stopped with 0.5 ml Na_2_CO 1M. OD_420_ and OD_550_ were measured. To obtain the activity (Miller) units, the following formula was applied: 1000×[(OD_420_ − (1,75 × OD_550_))/ t × 0,1 × OD_600_]

***Meta-analysis for SIRPβ1 (rs2209313).*** Participants in this study were obtained from multiple sources, including case-control summary statistics in the GR@ACE/DEGESCO^5^ cohort, in the International Genomics of Alzheimer’s Project (IGAP Stage I), the AD-by-proxy phenotype from the UK Biobank, the AddNeuroMed Study (ADDN) and the Neocodex–Murcia study. We performed a fixed-effects inverse-variance–weighted meta-analysis on the six independent studies using METAL software^6^.

*GR@ACE/DEGESCO:* The GR@ACE study recruited Alzheimer’s disease (AD) patients from Fundació ACE, Institut Català de Neurociències Aplicades (Catalonia, Spain), and control individuals from three centers: Fundació ACE (Barcelona, Spain), Valme University Hospital (Seville, Spain), and the Spanish National DNA Bank–Carlos III (University of Salamanca, Spain) (http://www.bancoadn.org). Additional cases and controls were obtained from dementia cohorts included in the Dementia Genetics Spanish Consortium (DEGESCO). At all sites, AD diagnosis was established by a multidisciplinary working group—including neurologists, neuropsychologists, and social workers—according to the DSM-IV criteria for dementia and the National Institute on Aging and Alzheimer’s Association’s (NIA–AA) 2011 guidelines for diagnosing AD. In our study, we considered as AD cases any individuals with dementia diagnosed with probable or possible AD at any point in their clinical course. Written informed consent was obtained from all the participants. The ethics and scientific committees have approved this research protocol (Acta 25/2016, Ethics Committee H., Clinic I Provincial, Barcelona, Spain).

*IGAP:* The GWAS summary results from the IGAP were downloaded from the National Institute on Aging Genetics of Alzheimer’s Disease Data Storage Site (NIAGADS, https://www.niagads.org/). Details on data generation and analyses by the IGAP have been previously described. In brief, the IGAP is a large study based upon genome-wide association using individuals of European ancestry. Stage 1 of the IGAP comprises 21,982 AD cases and 41,944 cognitively normal controls from four consortia: the Alzheimer Disease Genetics Consortium (ADGC), the European Alzheimer’s Disease Initiative (EADI), the Cohorts for Heart and Aging Research in Genomic Epidemiology (CHARGE) Consortium, and the Genetic and Environmental Risk in AD/Defining Genetic, Polygenic, and Environmental Risk for Alzheimer’s Disease (GERAD/PERADES) Consortium. Summary statistics are available for 11,480,632 variants, both genotyped and imputed (1000 Genomes phase 1, v3).

*UK Biobank*: UK Biobank data - including health, cognitive, and genetic data - was collected on over 500,000 individuals aged 37–73 years from across Great Britain (England, Wales, and Scotland) at the study baseline (2006–2010) (http://www.ukbiobank.ac.uk). Several groups have demonstrated the utility of self-report of parental history of AD for case ascertainment in GWAS (proxy–AD approach). For this study, we used the published summary statistics^7^. They included, after stringent QC, 314,278 unrelated individuals for whom AD information was available on at least one parent in the UK Biobank (https://datashare.is.ed.ac.uk/handle/10283/3364). In brief, the 27,696 participants whose mothers had dementia (maternal cases) were compared with the 260,980 participants whose mothers did not have dementia. Likewise, the 14,338 participants whose fathers had dementia (paternal cases) were compared with the 245,941 participants whose fathers did not have dementia. The phenotype of the parents is independent, and therefore, the estimates could be meta-analyzed. After analysis, the effect estimates were made comparable to a case-control setting. The data available comprises summary statistics of 7,794,553 SNPs imputed to the HRC reference panel (full panel).

*The Neocodex–Murcia study* includes 324 sporadic AD patients and 754 controls of unknown cognitive status from the Spanish general population collected by Neocodex. AD patients were diagnosed as having possible or probable AD in accordance with the NINCDS–ADRDA criteria.

*The AddNeuroMed Study (ADDN)* was a public–private partnership for biomarker discovery and replication in AD43–45. It was a multicenter study in Europe, with the first patient enrolled in January 2006 and the last in February 2008. The study protocol was planned for a baseline assessment visit, with follow-ups every three months for the first year and then annual visits that continued through 2013. The study enrolled a total of 258 AD, 257 MCI, and 266 controls, but not all had complete data at each assessment. In our study, we included 450 cases and 187 controls.

**Linkage disequilibrium between rs2209313 and the *SIRPβ1* intragenic duplication**

In order to evaluate the use of rs2209313 as a proxy for the genotyping of the duplication, we analyzed the linkage disequilibrium between both variants using 1,000G data. Briefly, 1,000G phased data including single nucleotide variants, indels and structural variants was retrieved for chromosome 20 from <http://ftp.1000genomes.ebi.ac.uk/vol1/ftp/data_collections/1000G_2504_high_coverage/working/20220422_3202_phased_SNV_INDEL_SV/> and <<http://ftp.1000genomes.ebi.ac.uk/vol1/ftp/data_collections/1000G_2504_high_coverage/working/20220422_3202_phased_SNV_INDEL_SV/>>. Bcftools v1.7 were used to retrieve genotype information for CEU, YRI and CHB population and variants rs2209313 and hgsv235481 located in chr20:1618496 and chr20:1580436, respectively. Pould R package was used to estimate LD between markers for the entire dataset (N= 310, D’= 0.963). Estimates for each populations were as follows: CEU= 0.969 , CHB=1, and YRI= 0.903.

**GR@CE series**

***Participants.*** The ACE biomarker research program included individuals evaluated consecutively at the ACE Alzheimer center Memory Clinic (Barcelona, Spain). All participants completed neurological, neuropsychological and social evaluations. A consensus diagnosis was assigned to each patient by a multidisciplinary team of neurologists, neuropsychologists, and social workers. All subjects were examined with the Mini-Mental State Examination (MMSE), Hachinski Ischemia Scale, and Clinical Dementia Rating (CDR) scale and a comprehensive neuropsychological battery of Ace Alzheimer center (N-BACE). Dementia was defined according to the DSM-V criteria. Mild cognitive impairment (MCI) was defined using Petersen´s criteria. Participants were selected based on their MCI diagnosis within the ACE Alzheimer Center biorepository (n=493).

**Extended References**
